# Predictors of Physician Awareness of the Periodontal Disease–Diabetes Association: A Cross-Sectional Study in Ghana

**DOI:** 10.64898/2026.04.08.26350446

**Authors:** Gloria A. M. Fiifi-Yankson, Edward Ohene-Marfo, Franklin N. Glozah, Edward Nordjo, Denise A. Mantey, Daniel Tormeti, Roma Garner, Josephine Sackeyfio

## Abstract

**Background:** Periodontal disease (PD) and diabetes mellitus (DM) have a well-established bidirectional relationship, affecting glycaemic control and chronic disease outcomes. However, the extent to which medical training supports physician awareness of this association remains unclear especially in resource-limited settings.

**Objective:** To assess exposure to oral health education and to identify predictors of awareness of PD-DM association among physicians.

**Methods:** A cross-sectional study was conducted among 146 physicians managing diabetic patients at a tertiary teaching hospital in Ghana. A structured questionnaire assessed exposure to oral health education, periodontal disease knowledge (score range 0–5), and awareness using a 5-item Likert scale (score range 5–25). Multivariable linear regression identified predictors of awareness.

**Results:** Although 62.1% reported exposure to oral health content during undergraduate training, 59.2% rated its quality as poor. Mean awareness score was 20.6 (SD=2.8). Awareness was independently predicted by years of professional experience (p < 0.001) and periodontal disease knowledge (p = 0.008), but not by structured oral health curriculum exposure.

**Conclusion:** Awareness of the PD-DM link was high but was not explained by formal educational exposure. Awareness appears to develop through knowledge of PD and professional experience, suggesting a gap between curricular exposure and competency.

## Introduction

Periodontal diseases (PD) are inflammatory oral conditions, defined as destruction of the supporting tissues of the teeth, namely the gingiva, the periodontal ligament, and the alveolar bone. Globally, PD is prevalent in about 90% of adults, with burden rising with age ^1,2^. Diabetes mellitus (DM) is a chronic condition characterized by dysfunction in the production and utilization of insulin, and it is on the rise with immense implications for the cost of healthcare if not tackled expeditiously ^3^. An estimated 1.1 % rise in diabetes prevalence is expected by 2045 globally, according to Lovic et al ^4^. Prevalence of diabetes is expected to increase by 156% in Africa alone by the year 2045 ^5^. Several chronic complications accompany long-term glucose dysregulation, including PD ^6^.

It has been largely recognized that a bidirectional link exists between PD and DM (both type 1 and type 2). PD exerts influence on glycaemic control whilst uncontrolled DM potentiates PD ^7,8^. A recent meta-analysis on the prevalence of periodontal diseases between diabetics and controls showed a higher prevalence of PD among diabetics (67.8%) in comparison to the non-diabetics (35.5%) ^9^, with a 3-fold increased odds of having periodontitis compared to non-diabetic subjects in another study ^10^. Non-surgical periodontal therapy has been shown to improve glycaemic control among individuals with diabetes ^11^.

Despite growing evidence supporting the PD–DM association, oral health integration within medical education remains inconsistent. Many physicians have reported receiving limited or no oral health education during training^12–14^. However, the absence of formal education may not uniformly translate into low awareness of concepts such as the PD-DM relationship.

Various guidelines for prevention and management of diabetes have recommended incorporation of oral health reviews and referrals into their regimen^15–17^. These reflect a broader shift towards integrated models of care in the management of chronic diseases such as diabetes. However, integrating oral health into medical practice remains limited globally, particularly in settings where there is a lack of oral health content in training programs, despite the growing body of evidence and guideline recommendations.

Awareness of the PD–DM association among physicians has been variably reported across settings. A study in 2022 showed that over one-third of primary care physicians and specialists were unaware of the link^18^. Awareness of the PD-DM association has also been reported to be higher in some populations(Taşdemir & Alkan, 2015; Panakhup et al., 2021) than in Africa ^20^. Longer years of practice and a higher level of the job rank in terms of professional experience may bring with it exposure to information regarding the association. A study in Jordan observed that specialists had higher awareness of this association compared to general practitioners^21^. A good understanding of periodontal disease may also increase a physician’s awareness of the PD-DM link. Chatzaki et al showed that physicians with good knowledge of periodontitis also had higher awareness of the relationship ^12^.

This variation suggests that awareness might be influenced by a combination of formal training, clinical exposure, and informal learning.

Few studies, have simultaneously examined the contributions of formal educational exposure, retained PD knowledge, and professional experience in shaping awareness, particularly in Sub-Saharan Africa. Ghana’s burden of oral diseases is gradually climbing ^22^, yet there is a limited dentist-to-population ratio ^23^. In these limited settings where physicians often serve as first contact for patients with undiagnosed oral conditions, their awareness is critical for integrated care. However, the extent to which awareness is driven by curriculum exposure versus experiential learning remains unclear.

Additionally, the majority of persons living with diabetes (PLWD) in Ghana have limited information on this link as observed by Broder et al ^24^. This makes it key for physicians to be aware of this relationship to facilitate holistic care for PLWD. The extent to which medical training equips physicians, especially in Ghana, to recognize and respond to these oral–systemic interactions currently remain unclear. Clarifying whether awareness is driven by curriculum exposure, knowledge acquisition, or professional maturation may help inform strategies for strengthening oral–systemic integration in medical training and during continuous professional development.

This study aimed to assess the level of awareness of the periodontal disease–diabetes association among physicians managing diabetic patients in the largest tertiary teaching hospital in Ghana. It also sought to evaluate the contributions of the oral health curriculum exposure, periodontal disease knowledge, and years of professional experience to awareness of the PD–DM relationship.

## Materials and Methods

### Study Design and Setting

This cross-sectional study was conducted among physicians managing patients with diabetes mellitus at Korle Bu Teaching Hospital, Accra, Ghana. The study is reported in accordance with the Strengthening the Reporting of Observational Studies in Epidemiology (STROBE) guidelines for cross-sectional studies.

Korle Bu Teaching Hospital is the largest tertiary referral centre and a major teaching hospital in Ghana. It serves as a training site for undergraduate and postgraduate medical education and provides specialized care for patients with chronic conditions, including diabetes mellitus.

Physicians in the Departments of Internal Medicine and Family Medicine were included due to their key role in diabetes management.

### Participants and Sampling

The study population comprised all physicians working within the departments of Internal Medicine and Family Medicine during the study period from September to October 2022. Physicians not engaged in active clinical patient care and newly recruited doctors were excluded.

A finite population correction was applied to calculate the required sample size from a population of 203 eligible physicians. Cochran’s formula was used with an assumed population proportion of 50%, a 5% margin of error, and 95% confidence level. A minimum sample size of 133 was calculated and adjusted for a 10% anticipated non-response rate, yielding an approximated sample of 146 physicians.

A multistage sampling was used in this study. First, stratified random sampling was used on the two strata corresponding to the Departments of Internal Medicine and Family Medicine, which collectively included 203 eligible physicians (139 in Internal Medicine and 64 in Family Medicine). Based on the total sample size of 146, the proportional allocation yielded target samples of approximately 100 physicians from Internal Medicine and 46 from Family Medicine. A simple random sample using a computer-generated random number sequence was then carried out to select the required number of physicians from each department.

Data collection took place on all clinic days within the study period, and data collectors received training to ensure standardized and accurate administration. Unique identification codes were assigned to participants to facilitate tracking and to prevent duplicate entries.

### Data Collection and Analysis

Data was collected using a structured self-administered questionnaire. Four main domains covered were:

1. **Sociodemographic characteristics** (age, sex, years of professional experience, department, job cadre, history of dental visits).
2. Exposure to oral health education during medical training**, including:**

a. Presence of oral health content in undergraduate curriculum (Yes/no)
b. Affiliation of training institution with a dental school (Yes/no)
c. Clinical dental exposure during training (Yes/no)
d. Self-rated quality of oral health education(good/fair/poor)
3. **Knowledge of periodontal disease** assessed using five multiple-choice items covering definition, etiology, early signs, and prevention (score range: 0–5).
4. **Awareness of the periodontal disease–diabetes association**, measured using a 5-item Likert scale (1 = strongly disagree to 5 = strongly agree; composite score range: 5–25). Items assessed recognition of PD as a complication of diabetes, increased periodontal risk in diabetes, the impact of periodontal health on glycaemic control, and potential benefits of periodontal therapy on glycaemic control.

Face validity of the questionnaire was established through review by specialists in periodontology and internal medicine. Revisions were made based on feedback before final administration.

Internal consistency of the 5-item awareness scale was evaluated using Cronbach’s alpha coefficient. A threshold of α ≥ 0.70 was considered acceptable for scale reliability.

Data were entered into Microsoft Excel and analysed using SPSS version 25 (IBM Corp., Armonk, NY, USA).

Descriptive statistics were used to summarize participant characteristics, educational exposure variables, knowledge scores, and awareness scores. Categorical variables were presented as frequencies and percentages. Continuous variables were presented as means and standard deviations.

Assumptions of linear regression were evaluated through inspection of standardized residual histograms, normal probability (P–P) plots, and residual scatterplots. No substantial deviations from normality, linearity, or homoscedasticity were observed. One-way ANOVA test were used to assess differences in mean awareness across physician characteristics.

Linear regression was done to identify predictors of PD-DM awareness among physicians. The variables included in the regression model were selected based on their theoretical relevance and reviewed literature. Statistical significance for this study was set at *p* < 0.05.

Ethical approval was obtained from the Institutional Review Board at the Korle Bu Teaching Hospital (ID NO. KBTH-STC 000140/2022). A written informed consent was obtained from each participant before the administration of the questionnaire. Participation was voluntary, and no compensation or incentives were given to participants in this study.

## Results

### Participant Characteristics

A total of 146 physicians participated in the study. Participants were drawn from the Departments of Internal Medicine and Family Medicine. Majority of participants were between 30 and 39 years of age (74.7%), and 56.2% were male (Table 1). Most respondents had between 6 to 10 years of professional experience (43.2%), and over half were residents (52.7%).

**Table 1.**
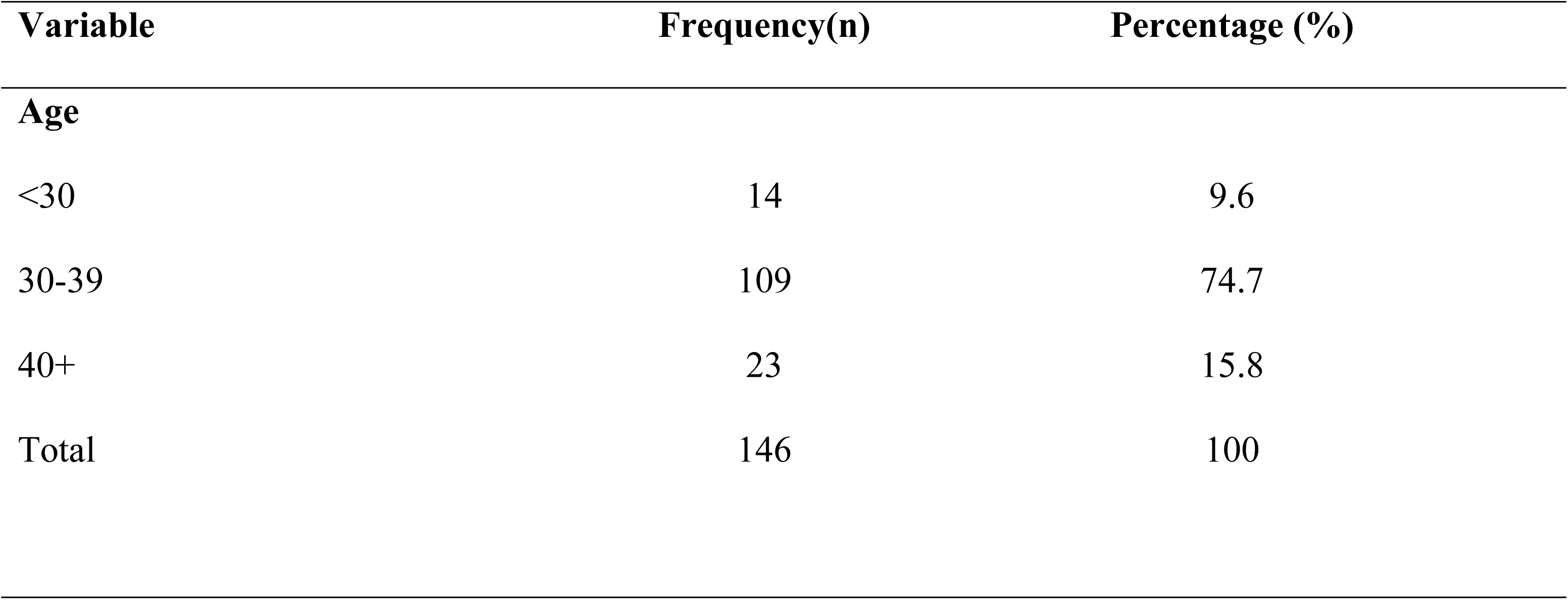
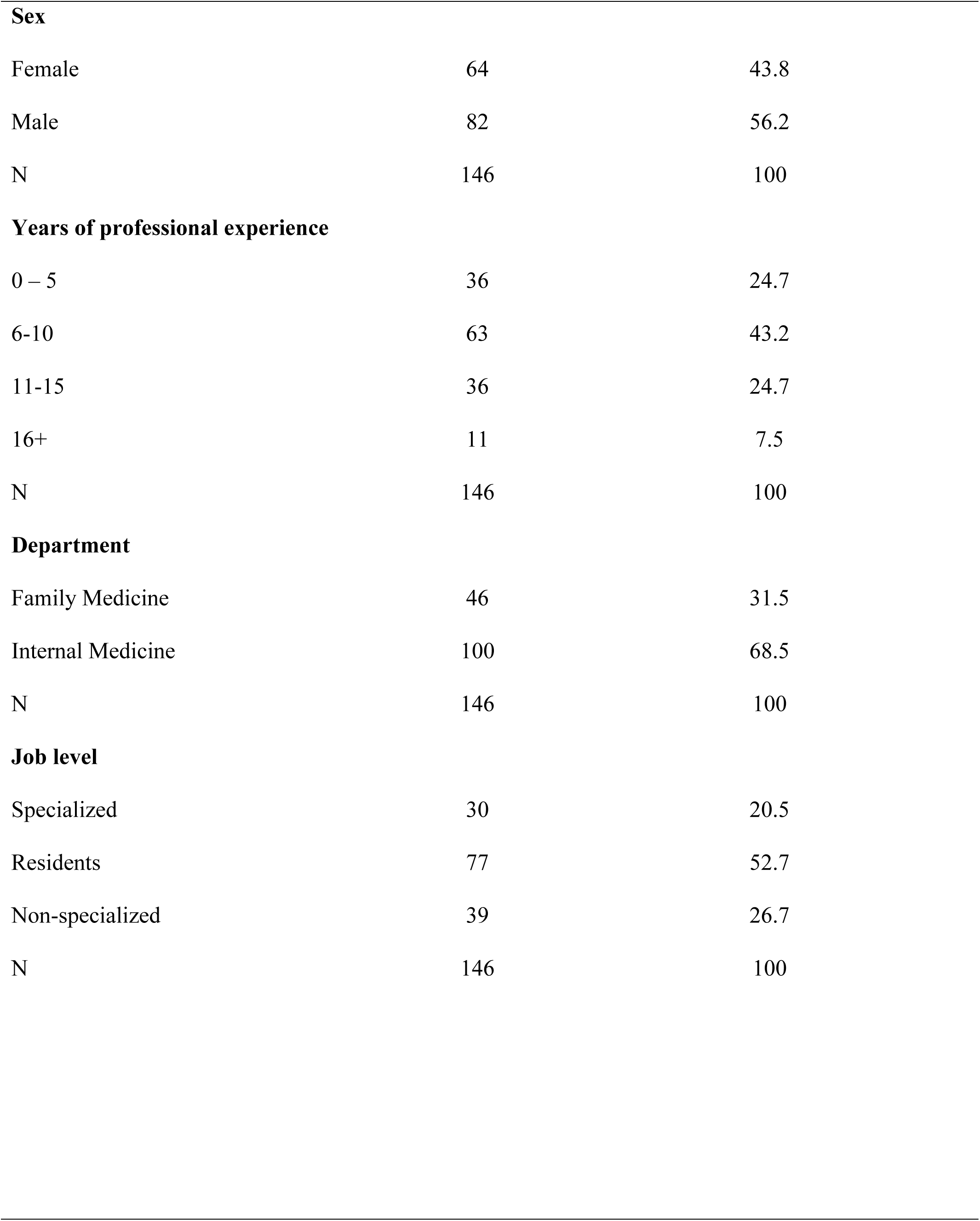
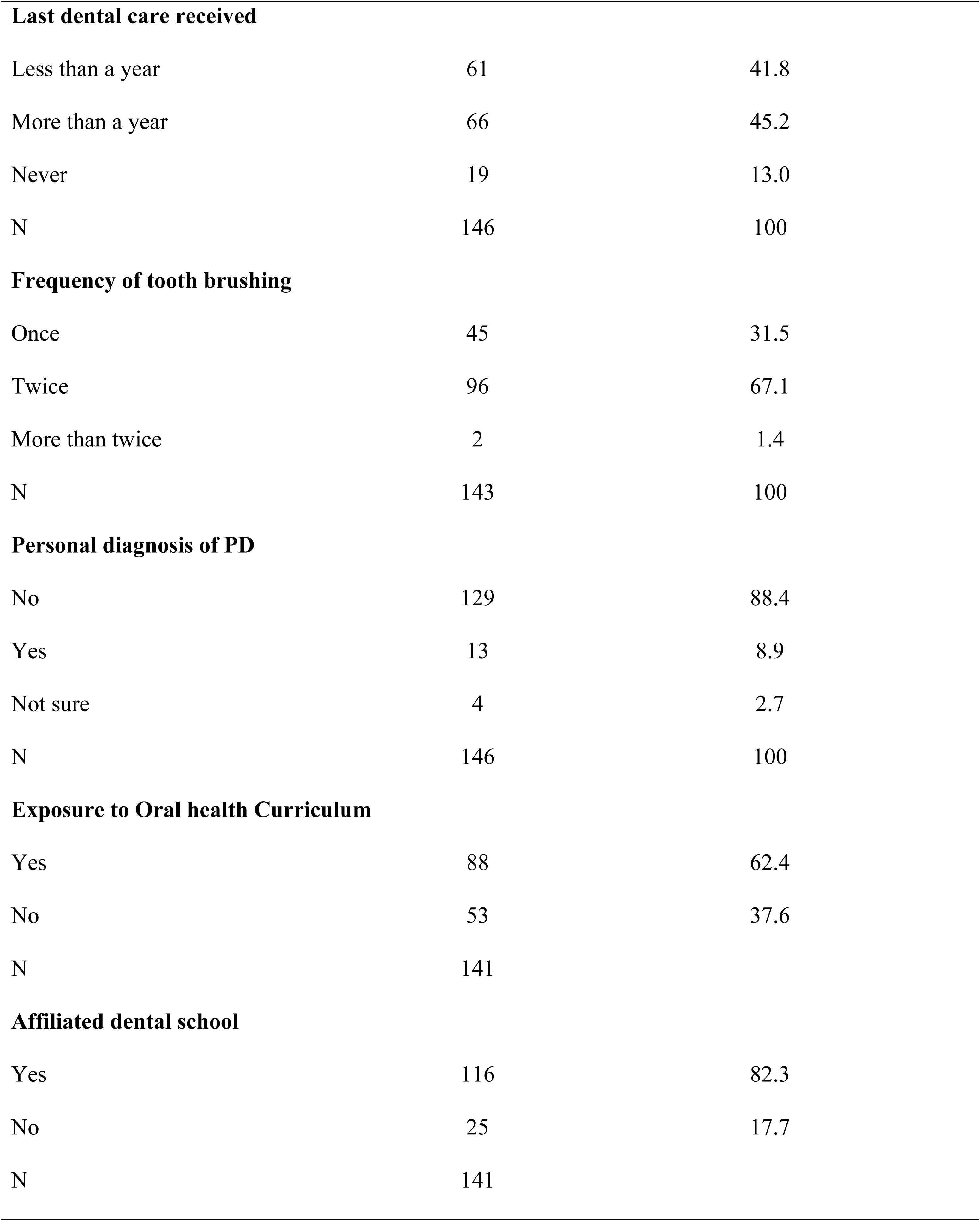
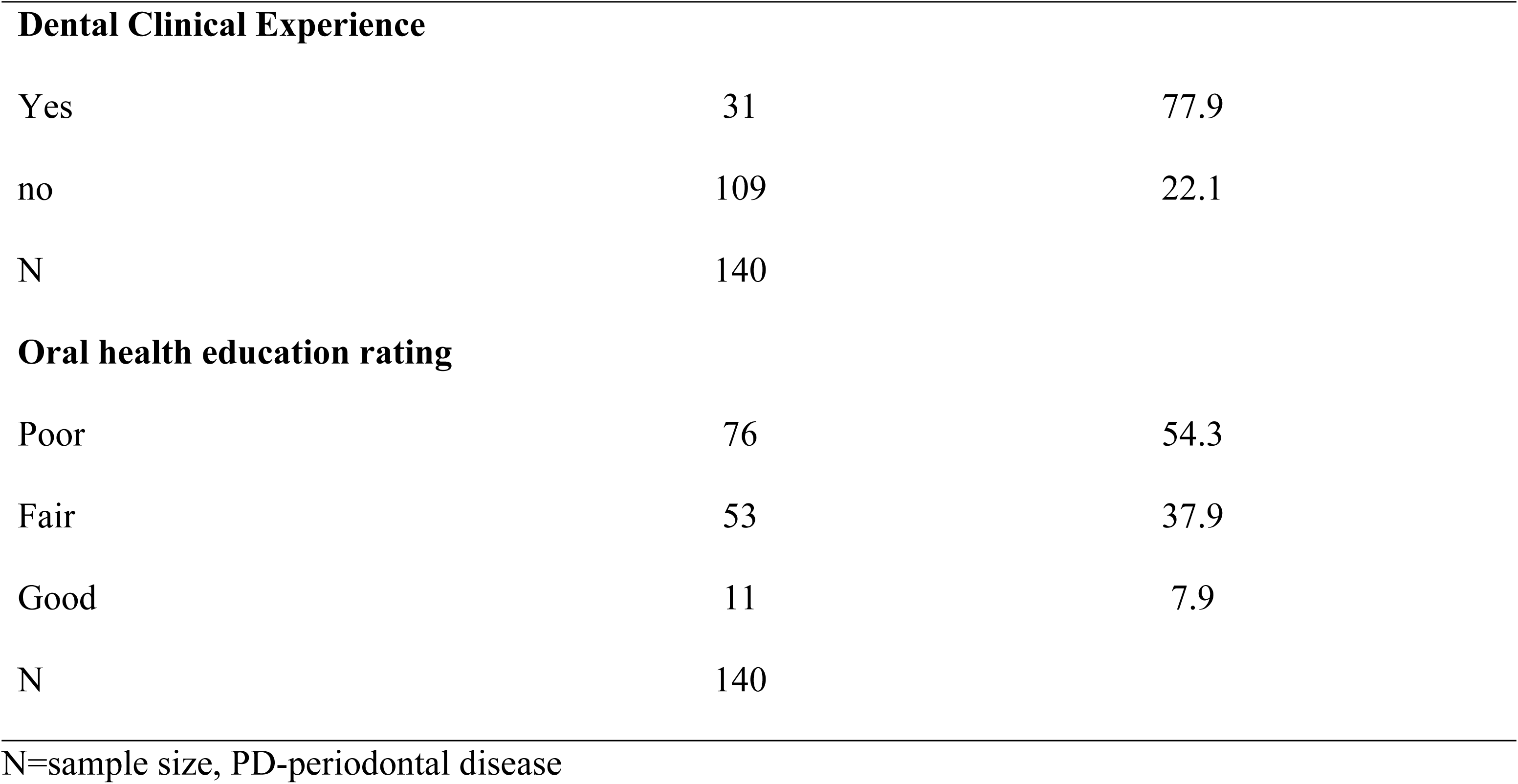
Distribution of Sociodemographic Characteristics.

| Variable | Frequency(n) | Percentage (%) |
| --- | --- | --- |
| **Age** |  |  |
| <30 | 14 | 9.6 |
| 30-39 | 109 | 74.7 |
| 40+ | 23 | 15.8 |
| Total | 146 | 100 |
| **Sex** |  |  |
| Female | 64 | 43.8 |
| Male | 82 | 56.2 |
| N | 146 | 100 |
| **Years of professional experience** |  |  |
| 0 – 5 | 36 | 24.7 |
| 6-10 | 63 | 43.2 |
| 11-15 | 36 | 24.7 |
| 16+ | 11 | 7.5 |
| N | 146 | 100 |
| **Department** |  |  |
| Family Medicine | 46 | 31.5 |
| Internal Medicine | 100 | 68.5 |
| N | 146 | 100 |
| **Job level** |  |  |
| Specialized | 30 | 20.5 |
| Residents | 77 | 52.7 |
| Non-specialized | 39 | 26.7 |
| N | 146 | 100 |
| **Last dental care received** |  |  |
| Less than a year | 61 | 41.8 |
| More than a year | 66 | 45.2 |
| Never | 19 | 13.0 |
| N | 146 | 100 |
| **Frequency of tooth brushing** |  |  |
| Once | 45 | 31.5 |
| Twice | 96 | 67.1 |
| More than twice | 2 | 1.4 |
| N | 143 | 100 |
| **Personal diagnosis of PD** |  |  |
| No | 129 | 88.4 |
| Yes | 13 | 8.9 |
| Not sure | 4 | 2.7 |
| N | 146 | 100 |
| **Exposure to Oral health Curriculum** |  |  |
| Yes | 88 | 62.4 |
| No | 53 | 37.6 |
| N | 141 |  |
| **Affiliated dental school** |  |  |
| Yes | 116 | 82.3 |
| No | 25 | 17.7 |
| N | 141 |  |
| **Dental Clinical Experience** |  |  |
| Yes | 31 | 77.9 |
| no | 109 | 22.1 |
| N | 140 |  |
| **Oral health education rating** |  |  |
| Poor | 76 | 54.3 |
| Fair | 53 | 37.9 |
| Good | 11 | 7.9 |
| N | 140 |  |

N=sample size, PD-periodontal disease

Most physicians (62.4%) reported receiving some oral health content during undergraduate medical training. However, 54.3% rated the quality of oral health education to be poor.

Although 82.3% of respondents were trained at institutions affiliated with a dental school, only 22.1% reported any form of clinical dental exposure during medical training. This suggests that while theoretical exposure was common, structured clinical exposure to oral health was limited. Additional demographic characteristics are presented in Table 1.

### Other Sources of Information on the PD–DM Association

When physicians were asked to identify their primary sources of information regarding the periodontal disease–diabetes association, learning pathways appeared largely informal and practice-based rather than curriculum-driven (Figure 1).

**Figure 1.**
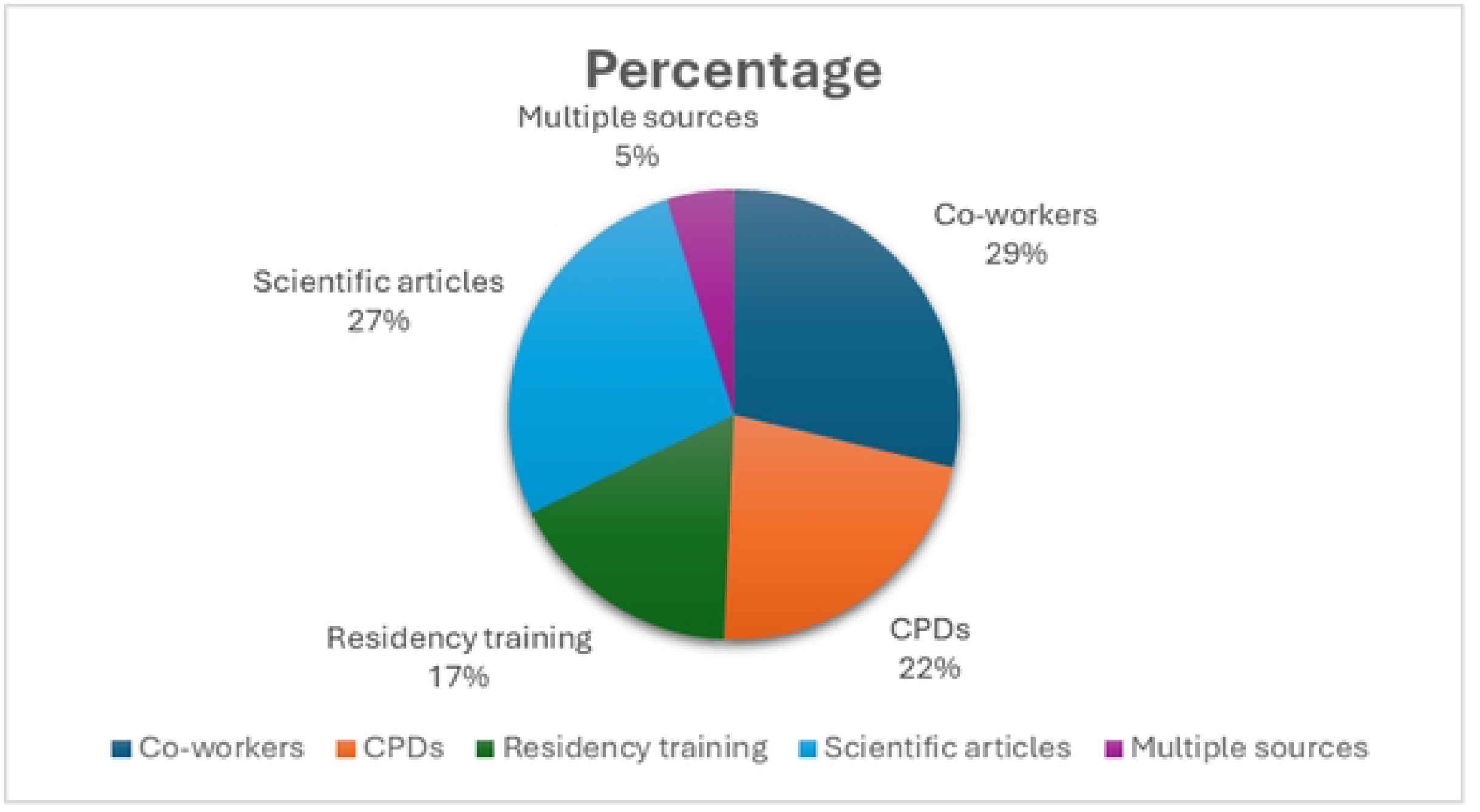
Distribution of Other Sources of Information on the PD–DM Association.

Co-workers were the most frequently cited source (29%), followed closely by scientific articles (27%) and continuing professional development (CPD) activities (22%). Residency training accounted for 17% of responses, while only 5% reported drawing from multiple sources.

### Knowledge of Periodontal Disease Scores and Awareness of PD-DM Scores

The mean periodontal disease knowledge score was 3.6 (SD = 0.8) on a 0–5 scale. The five-item awareness scale demonstrated good internal consistency (Cronbach’s α = 0.84). The summed awareness score ranged from 5 to 25, with a mean of 20.6 (SD = 2.8), indicating generally high levels of awareness.

### Awareness of Periodontal Disease–Diabetes link Across Physician Characteristics

Significant differences in mean awareness were observed across age (F (2,134) = 6.43, p = 0.002, η² = 0.088), job level (F (2,134) = 6.69, p = 0.002, η² = 0.091), and years of professional experience (F (3,133) = 5.12, p = 0.002, η² = 0.104). Awareness increased with age and experience (Table 2). No significant differences were observed across sex, department, or educational exposure variables. Thus, awareness differences were driven primarily by age, job levels, and years of experience, whereas other demographic factors did not show meaningful variation.

**Table 2.**
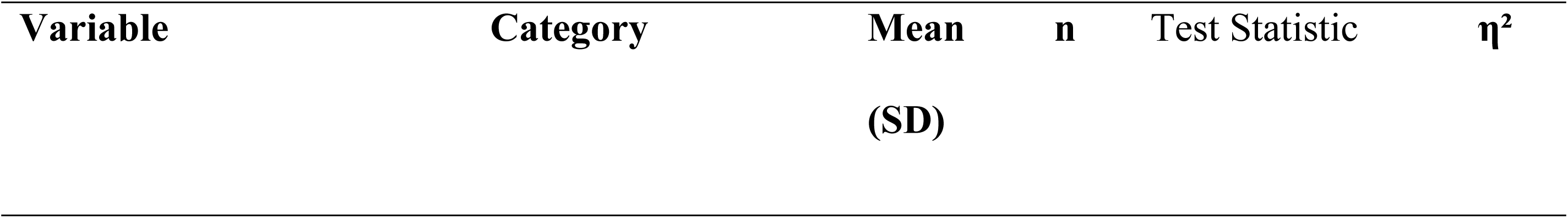
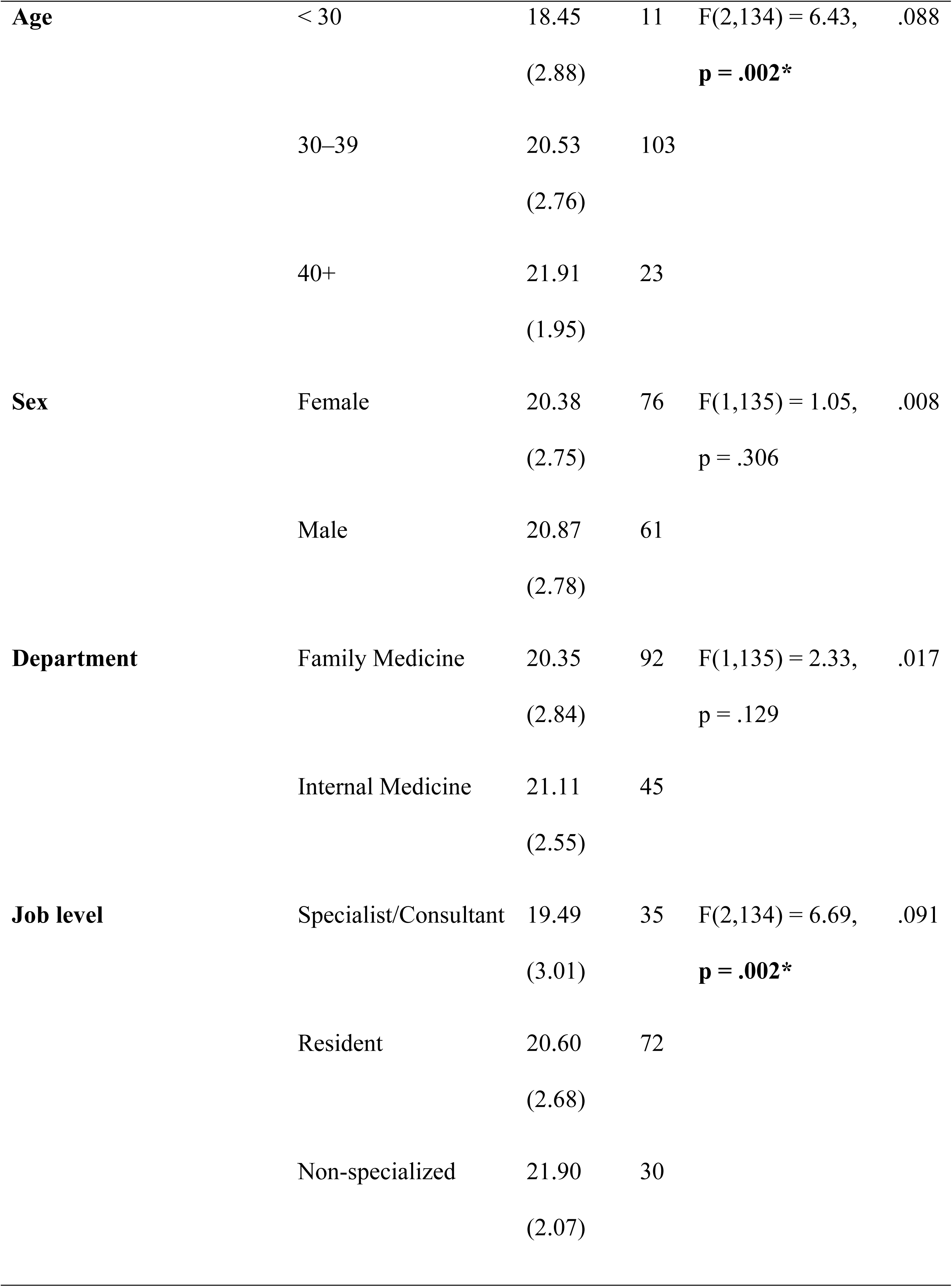
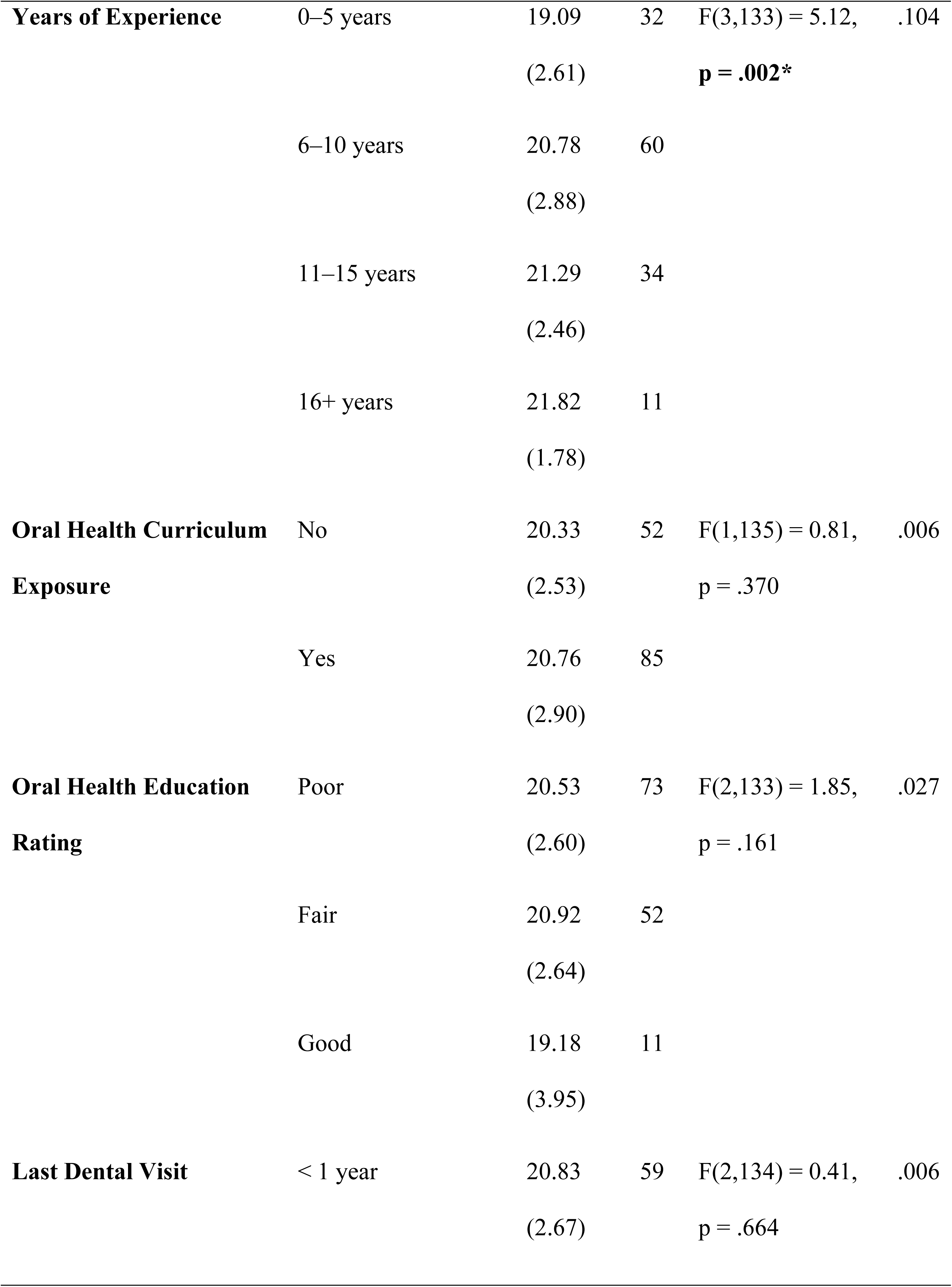
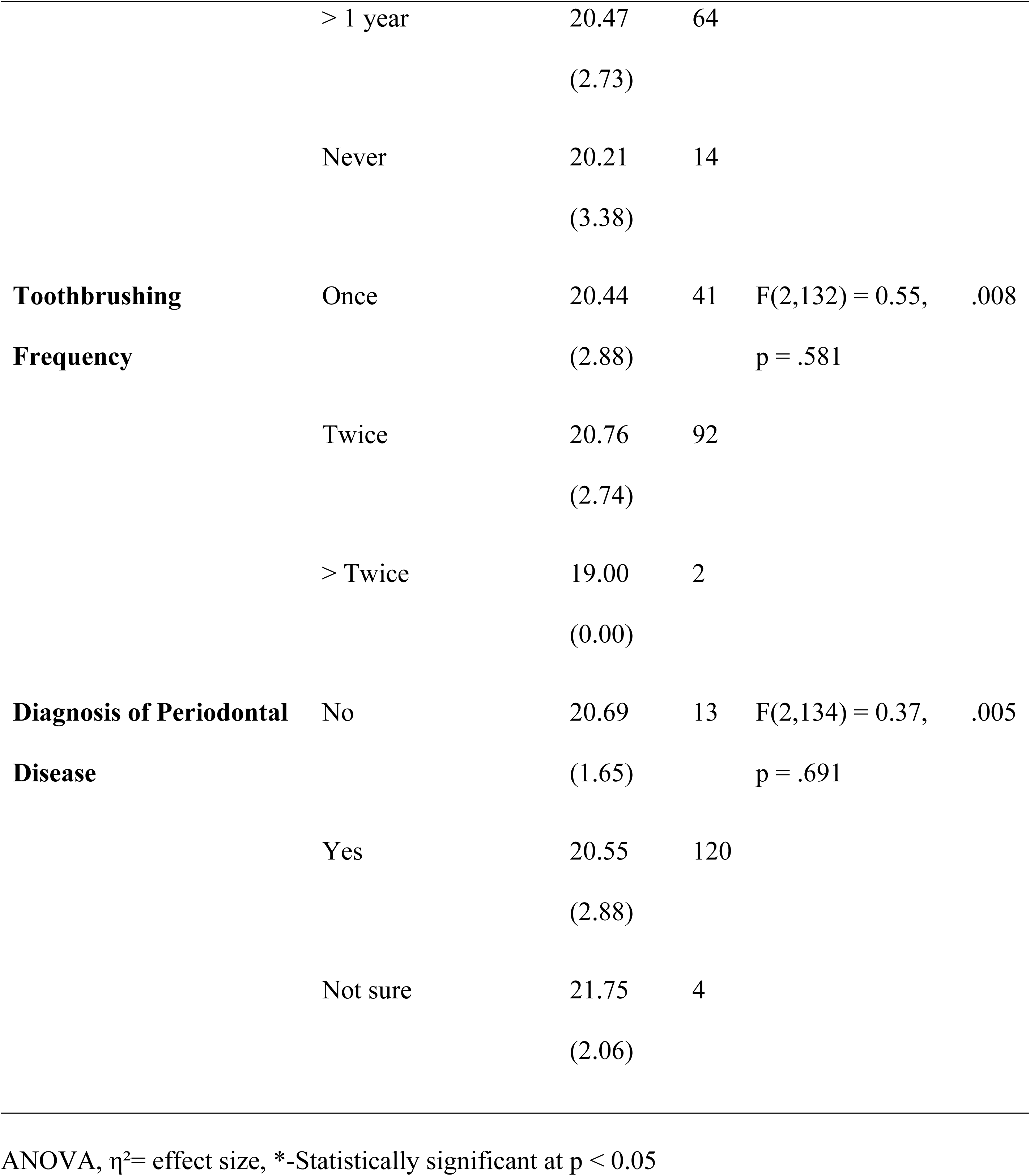
Bivariate Associations Between Physician Characteristics and Awareness.

| Variable | Category | Mean<br>(SD) | n | Test Statistic | $\eta^2$ |
| --- | --- | --- | --- | --- | --- |
| **Age** | < 30 | 18.45<br>(2.88) | 11 | F(2,134) = 6.43,<br>**p = .002\*** | .088 |
|  | 30–39 | 20.53<br>(2.76) | 103 |  |  |
|  | 40+ | 21.91<br>(1.95) | 23 |  |  |
| **Sex** | Female | 20.38<br>(2.75) | 76 | F(1,135) = 1.05,<br>p = .306 | .008 |
|  | Male | 20.87<br>(2.78) | 61 |  |  |
| **Department** | Family Medicine | 20.35<br>(2.84) | 92 | F(1,135) = 2.33,<br>p = .129 | .017 |
|  | Internal Medicine | 21.11<br>(2.55) | 45 |  |  |
| **Job level** | Specialist/Consultant | 19.49<br>(3.01) | 35 | F(2,134) = 6.69,<br>**p = .002\*** | .091 |
|  | Resident | 20.60<br>(2.68) | 72 |  |  |
|  | Non-specialized | 21.90<br>(2.07) | 30 |  |  |
| **Years of Experience** | 0–5 years | 19.09<br>(2.61) | 32 | F(3,133) = 5.12,<br>**p = .002\*** | .104 |
|  | 6–10 years | 20.78<br>(2.88) | 60 |  |  |
|  | 11–15 years | 21.29<br>(2.46) | 34 |  |  |
|  | 16+ years | 21.82<br>(1.78) | 11 |  |  |
| **Oral Health Curriculum Exposure** | No | 20.33<br>(2.53) | 52 | F(1,135) = 0.81,<br>p = .370 | .006 |
|  | Yes | 20.76<br>(2.90) | 85 |  |  |
| **Oral Health Education Rating** | Poor | 20.53<br>(2.60) | 73 | F(2,133) = 1.85,<br>p = .161 | .027 |
|  | Fair | 20.92<br>(2.64) | 52 |  |  |
|  | Good | 19.18<br>(3.95) | 11 |  |  |
| **Last Dental Visit** | < 1 year | 20.83<br>(2.67) | 59 | F(2,134) = 0.41,<br>p = .664 | .006 |
|  | > 1 year | 20.47<br>(2.73) | 64 |  |  |
|  | Never | 20.21<br>(3.38) | 14 |  |  |
| **Toothbrushing**<br>**Frequency** | Once | 20.44<br>(2.88) | 41 | F(2,132) = 0.55,<br>p = .581 | .008 |
|  | Twice | 20.76<br>(2.74) | 92 |  |  |
|  | > Twice | 19.00<br>(0.00) | 2 |  |  |
| **Diagnosis of Periodontal**<br>**Disease** | No | 20.69<br>(1.65) | 13 | F(2,134) = 0.37,<br>p = .691 | .005 |
|  | Yes | 20.55<br>(2.88) | 120 |  |  |
|  | Not sure | 21.75<br>(2.06) | 4 |  |  |

ANOVA, $\eta^2$ = effect size, \*-Statistically significant at $p < 0.05$

### Predictors of Awareness of the Periodontal Disease–Diabetes Association

A multivariable linear regression analysis (n = 136) was conducted to evaluate whether oral health curriculum exposure predicted awareness (Table 3). The model was adjusted for years of professional experience and periodontal disease knowledge. The overall model was statistically significant (F (3,132) = 6.377, p < 0.001) and explained 12.7% of the variance in awareness (R² = 0.127).

**Table 3.** Multiple Linear Regression Predicting Awareness (n = 136)

| Predictor | B | SE | $\beta$ | t | p-value |
| --- | --- | --- | --- | --- | --- |
| Curriculum exposure (Yes/No) | 0.273 | 0.466 | 0.048 | 0.587 | 0.558 |
| Periodontal disease knowledge | 0.520 | 0.242 | 0.175 | 2.151 | 0.033* |
| Years of professional experience | 0.947 | 0.260 | 0.298 | 3.644 | <0.001* |
| Constant | 16.599 | 1.044 | — | 15.904 | <0.001 |

Model statistics: $F(3,132) = 6.377$ , $p < 0.001$ ; $R^2 = 0.127$ , \*Statistically significant at $p < 0.05$

Years of professional experience (B = 0.947, p < 0.001) and periodontal disease knowledge (B = 0.520, p = 0.033) were independent predictors of awareness. Oral health curriculum exposure was not significantly associated with awareness after adjustment (p = 0.558).

## Discussion

This study examined whether structured oral health training exposure during medical education translated into awareness of the bidirectional association between periodontal disease and diabetes among physicians managing diabetic patients. Although most physicians reported some exposure to oral health content during undergraduate training, with many trained in institutions affiliated with dental schools, more than half rated that exposure as poor. Second, overall awareness of the PD-DM link was high. A multivariable analysis showed that physician awareness of the periodontal disease–diabetes association was independently associated with years of professional experience and periodontal disease knowledge, but not with reported oral health curriculum exposure. These findings position the PD–DM relationship not as a knowledge gap, but as an implementation and competency gap within medical education.

The relatively high awareness (20.6 out of a pos 25), observed in this cohort is consistent with findings from studies conducted in Europe, North America, and parts of Asia, where physicians are increasingly recognizing the association between diabetes and periodontal disease ^18,25–27^.

This is in contrast with variable awareness levels in other low- and middle-income countries such as Nigeria ^20^. This may be influenced by differences in training structure and continuing professional development opportunities. It may also reflect increasing global dissemination of evidence regarding oral–systemic interactions, as well as exposure to scientific literature and peer discussions. In this study, many physicians reported obtaining knowledge through professional interactions and self-directed reading. This aligns with literature suggesting that continuing medical education and informal knowledge networks contribute substantially to clinical awareness (McMahon, 2025).

The higher exposure to oral health content among physicians in this study contrasts with that from some international studies, which reported limited exposure to oral health education during medical training. Grocock et al. (2019) observed that many doctors reported minimal prior oral health instruction and limited confidence in managing oral conditions in the UK^28^. Little to no oral information during training was also reported among Swiss endocrinologists and general practitioners^12^. Morel et al. (2022) observed increased student-reported comfort in performing oral examinations following participation in a structured oral health clerkship^29^. This dissatisfaction was also observed by the majority of program directors in family residency training programs in the US, despite being mandated to include oral health education^30^. Silk also observed high variability in oral content within the mandated oral health content in family medicine training. This aligns with the low proportion of participants in our study who identified residency training as a source of oral health knowledge. These findings suggest that oral health education remains an area of perceived weakness within medical training programs, including specialty training.

Exposure to oral health curriculum did not predict awareness of PD-DM relationship in this study. This indicates that oral health may not be consistently integrated within broader medicine curricula to an equal and appreciable degree across settings or exposure may have occurred too long ago to have had an impact. As such, physicians will more likely have to depend on other sources for awareness about the PD-DM link.

Participants in this study reported co-workers and continuous professional education as frequent sources of oral health information. This shows that interprofessional education strategies can improve PD-DM awareness even when the opportunity is missed during initial medical training.

Positive evaluations of interprofessional education sessions have been reported by several studies. An increase in Readiness for Interprofessional Learning Scale (RIPLS) was reported in Canada following an interprofessional continuing education in oral-systemic health^31^. Similar positive changes in knowledge of other healthcare professionals related to DM management were also observed in a systematic review ^32^. These suggest that when oral health education is intentionally designed and integrated, improvements in knowledge and confidence may be achievable.

The significance of periodontal disease knowledge highlights the distinction between exposure and retained understanding. While exposure reflects opportunity to learn, knowledge reflects internalized understanding of domain-specific content (periodontal disease). The positive association between knowledge and awareness is consistent with educational theory and prior studies demonstrating that physicians with greater oral health knowledge are more likely to recognize systemic implications of periodontal disease. The findings suggest that strengthening periodontal disease knowledge may be key to sustaining physician awareness of oral–systemic links.

Years of professional experience were strongly predicted by awareness (β = 0.298). Physicians with greater experience demonstrated higher awareness even after adjustment for knowledge and curriculum exposure. In bivariate analysis, age and job level were also associated with awareness; however, these associations likely reflect overlapping constructs of professional maturation. A study in Kuwait also demonstrated higher awareness was associated with older physicians^33^. The findings from this study and other studies with similar associations may reflect cumulative clinical exposure to patients with diabetes, experiential learning, and repeated encounters with systemic complications that reinforce oral–systemic connections. However, this contrasts with findings from Hong Kong where experience was not associated with awareness^27^,possibly reflecting differences in curriculum structure and sources of knowledge. Tse et al reported undergraduate medical curriculum as their top source of oral-systemic information, which may be a reason for experience not making a difference.

The findings from this research point toward a curriculum–competency gap. The presence of oral health content within medical curricula does not necessarily ensure durable competency outcomes. In this study, awareness appeared more strongly associated with professional maturation than with reported educational exposure. If oral–systemic integration is to become a routine part of diabetes care, medical education may need to move beyond isolated didactic inclusion of oral health topics. Structured interprofessional learning experiences, case-based integration of oral complications within chronic disease modules, and competency-based assessments may be necessary to ensure that PD–DM connections are not merely introduced but internalized. Residency training may represent a critical opportunity for reinforcing these concepts during formative professional years.

This study contributes to a limited body of literature examining PD-DM awareness among physicians involved in DM care in sub-Saharan Africa. Its strengths included the use of a reliable composite awareness measure and multivariable modelling to distinguish between structured training and experiential influences.

In conclusion, although physicians demonstrated high PD-DM awareness, it was not independently predicted by structured oral health education exposure. Instead, awareness appeared to be shaped by professional seniority and periodontal disease knowledge. These findings suggest that current medical curricula may not consistently translate into sustained competency. Intentional competency-based integration and interprofessional approaches may be necessary to ensure consistent preparation of physicians for integrated chronic disease management.

## Study Limitations

Limitations included reliance on self-reported awareness and educational exposure, which may be subject to recall bias, and the lack of assessment of actual clinical behaviors or practices. The cross-sectional design further limits causal inference of findings.

## Acknowledgements

1. Administrative staff of departments of Internal medicine and Family medicine, Korle Bu Teaching Hospital, Accra, Ghana
2. John Tetteh, University of Manchester, Manchester, United Kingdom.
3. Dr Papa Kwesi Fiifi-Yankson, University of Ghana Medical Center, Accra, Ghana

## DECLARATIONS

### Ethics approval and consent to participate

The study received approval from the Ethical & Protocol Review Board of the Korle Bu Teaching Hospital, Accra, Ghana. The study protocol was reviewed and approved under approval identification number KBTH-STC 000140/2022.

The study was conducted in accordance with the ethical principles of the Declaration of Helsinki. Written informed consent was obtained from all participants prior to participation.

### Data Availability Statement

The dataset supporting this study is publicly available at Zenodo (DOI: 10.5281/zenodo.19407039). This dataset is used in multiple analyses addressing distinct research questions.

### Author Contributions

Conceptualization: Gloria A.M. Fiifi-Yankson, Franklin Glozah, Daniel Tormeti

Data curation: Gloria A.M. Fiifi-Yankson

Formal analysis: Gloria A.M. Fiifi-Yankson, Edward Ohene-Marfo

Funding acquisition: Gloria A.M. Fiifi-Yankson

Investigation: Gloria A.M. Fiifi-Yankson, Denise A. Mantey, Edward Nordjo

Methodology: Gloria A.M. Fiifi-Yankson, Edward Ohene-Marfo

Project administration: Gloria A.M. Fiifi-Yankson, Denise A. Mantey

Resources: Gloria A.M. Fiifi-Yankson

Software: Gloria A.M. Fiifi-Yankson, Edward Ohene-Marfo

Supervision: Gloria A.M. Fiifi-Yankson, Franklin Glozah

Validation: Gloria A.M. Fiifi-Yankson, Josephine Sackeyfio.

Visualization: Gloria A.M. Fiifi-Yankson, Roma Garner

Writing - original draft: Gloria A.M. Fiifi-Yankson, Edward Ohene-Marfo

Writing - review & editing: Gloria A.M. Fiifi-Yankson, Edward Ohene-Marfo, Roma Garner, Edward Nordjo, Josephine Sackeyfio.

